# Does regional tissue oxygenation vary in infants transported by road and air. A prospective observational within-subject analysis of regional oxygenation measured by near infrared spectroscopy in infants transported in Western Australia

**DOI:** 10.1101/2022.08.22.22278456

**Authors:** M. O’Dea, M. Saito-Benz, G. Tremblay, R. Trawber, S. Resnick, J. Tan, J. Davis

## Abstract

**Objective:** Infants in Western Australia who are unwell frequently require air retrieval for tertiary care. Travelling at altitude may worsen hypoxia.

**Design, Setting and Patients:** We examined tissue oxygenation using near infrared spectroscopy (NIRS) during retrieval by road and air. Study design was prospective and observational comparing cerebral/mesenteric oxygenation (crS0_2_ and mS0_2_) and fractional tissue extraction (cFTOE and mFTOE) in newborns transported by fixed-wing aircraft and road ambulance. Transport by fixed-wing aircraft was a combination of by road and by air. Transport by ambulance was by road only. Primary outcome measure was a within-subjects comparison of oxygenation and tissue oxygen extraction (flight vs. road transit). Measurements in those transported by road were made as control (first and third quarter of road transit).

**Results:** There were 24 infants transported by air and 31 by road. The median (interquartile range) gestations and weights were similar [39+1(31 – 41^+3^) vs. 40+1(35^+3^ – 43) wk; 3.3 (1.8 – 4.95) vs. 3.2 (2.2 – 4.2) Kg]. Most were treated for respiratory disease. There was no difference in SpO2, respiratory support or haemoglobin between groups. During transport by fixed-wing aircraft, infants had lower crS0_2_ (75.3 (6.2) vs 77.9 (6.2); p=<0.0001) and mS0_2_ (69.3 (16.6) vs. 74.2 (11.6); p=<0.01) and increased cFTOE (0.18 (0.07) vs. 0.21 (0.07), p<0.001) and mFTOE (0.22 (0.12) vs. 0.28 (0.17); P=0.006) than when travelling by road. There was no difference in the control group.

**Conclusion:** Newborn infants who travelled by air had lower cerebral saturations and greater oxygen tissue extraction when travelling by air. The pathogenesis and impact of these findings need further exploration.

- **What is already known on this topic** *- Neonatal transport is not without risk and travelling at altitude may worsen cerebral hypoxia*.
- **What this study adds –** *NIRS monitoring during neonatal transport is feasible and air travel demonstrated lower regional saturation and increased oxygen extraction*
- **How this study might affect research, practice or policy –***NIRS monitoring as an adjunctive tool may contribute to safer neonatal transfers*

## Background and Aim

Regionalization and centralization of neonatal services requires a network of communication, education and transport^1^. Western Australia (WA) is the largest state in Australia and has an area of 2.6 million Km^2^. Some infants require transfer up to 2200 Km for tertiary neonatal care centralized in Perth, the capital city. The newborn emergency transport service of Western Australia (NETS WA) transports approximately 400 infants per year by air (Total transports = 1280 in 2021).

Aircrafts cruise between 25,000 and 40,000 feet (7620 - 12192 m) above sea level for reduced turbulence and fuel economy. At this altitude, there is insufficient oxygen to sustain life, so aircraft cabins are pressurized between 5000 to 8000 feet. At 8000 feet, the atmospheric pressure is 75 kPa (565 mm Hg), and an atmospheric partial pressure of oxygen (pO_2_) of 15.7 kPa (18 mmHg). This is equivalent to 15-16% ambient oxygen available at sea level (21%) ^2^.

Near infrared spectroscopy (NIRS) is a non-invasive technique for measuring tissue oxygenation and haemodynamics and may detect hypoxaemia in the brain and other vital organs before changes in pulse oximetry readings are obvious^3^. NIRS functions due to the transparency of biological tissues to light in the near infrared part of the spectrum (700-1000 nm) and subsequent absorption by oxygenated haemoglobin and deoxygenated haemoglobin in the cerebral blood vessels which are within the near infrared light beam^4^. Absorption changes in near infrared light can then be converted into concentration changes of oxygenated haemoglobin and deoxygenated haemoglobin^4^.

The impact of air transport on tissue oxygenation on newborn infants is relatively unknown. The aim of this study was to examine changes in cerebral and regional (mesenteric) oxygenation, measured by NIRS, in infants who required transport by air soon after birth.

## Methods

This study was a prospective observational and within subject analysis comparing cerebral and mesenteric regional oxygenation, measured by NIRS, during neonatal air and road transport. Ethical approval from the Child and Adolescent Service Research Ethics Committee (EC00268), and informed parental consent was obtained. There were no predefined selection criteria and all transported were eligible for NIRS monitoring by road and air transport. This research received no specific grant from any funding agency in the public, commercial or not-for-profit sectors.

For infants who required air transport (distance >150 km), travel from referral hospital to the aircraft is by road ambulance, then by aircraft to an airstrip near the receiving hospital and by road ambulance to receiving hospital. For shorter distances, infants who require road transport are only moved by road ambulance. For air transport infants were transported in a Pilatus PC12 (Stans, Switzerland) single engine turboprop fixed-wing aircraft which reaches a maximum cruising altitude of 7620 m (25,000 feet) and is pressurized to approximately 2400 m (8000 feet). Flights were coordinated by the Royal Flying Doctors Service (RFDS) Western Operations. All were transported using the Mansell Neocot™ (Toowoomba, QLD, Australia) which incorporates equipment essential to provide intensive care management during transport.

NIRS measurements were taken using the Equinox SenSmart X-100 and (Nonin Medical, MN, USA) at a sampling rate of 0.25 Hz. Sensors were placed on the forehead and right lower quadrant of the abdomen to record cerebral (crSO_2_) and mesenteric (mrSO_2_) tissue oxygenation respectively. Peripheral arterial saturation (SpO_2_) was monitored concurrently to allow calculation of cerebral and mesenteric fractional tissue oxygen extraction (FTOE = (SpO_2_ – rSO_2_)/SpO_2_). Regional FTOE gives an estimate of the balance between local oxygen delivery and consumption^4^. For patients being transferred by aircraft, NIRS monitoring was commenced at least 30 minutes prior to take-off and continuing after landing allowed observation and comparison of cerebral oxygenation during air flight and ground retrieval (altitude vs. to sea level).

A within subject analysis was performed of crSO_2_, mrSO_2_ and FTOE on the ground and at maximum altitude for infants retrieved by air and between the 1^st^ and 3^st^ quarter of road transportation. Demographic data (depending on the distribution) was compared by parametric or non-parametric analysis of central tendency for continuous variables (t test or Mann Whitney U test) or Chi Squared for proportional data. Paired t-test was used for comparison of regional oxygenation and oxygen extraction data. A p value ≤ 0.05 was statistically significant throughout.

## Results

A total of 55 infants were included, 24 infants transported by air and 31 by road Most infants transported were born at term (>37 weeks completed weeks of gestation) and transported on the first day of life. There was a greater proportion of male infants in both groups. Demographic data is presented in Table 1. Respiratory pathology made up the highest number of transfers in both groups. For infants transported by air, 13/24 (54%) required respiratory support, three were ventilated. For infants transported by road, 18/31 (58%) required respiratory support, none were ventilated. Peripheral oxygen saturation was similar between the groups. The reason for retrieval, respiratory support and physiology data are also presented in Table 1.

**Table 1:** Demographics of the infants retrieved by air and retrieved by road. Continuous variables are expressed as median (interquartile range; IQR), categorical variables are expressed as number (percentage). Infants transported by fixed-wing aircraft were transferred by road and air. Infants transported road ambulance were only transported by road.

| Parameter | Air (n=24) | Road (n=31) | p |
| --- | --- | --- | --- |
| Gestation at birth (weeks) | 38+5 (31 – 41<sup>+2</sup>) | 39+2 (33<sup>+4</sup> – 41<sup>+2</sup>) | 0.24 |
| Day of life transferred | 1 (0 – 1) | 1 (0 – 4) | 0.20 |
| Birth Weight (g) | 3310 (1800 – 4950) | 3206 (2220 – 4220) | 0.49 |
| Male sex | 17 (70.8%) | 21 (67.7%) | 0.81 |
| **Reason for retrieval** |  |  |  |
| Respiratory | 10 (41.7%) | 14 (45.2%) | 0.96 |
| Non-respiratory | 14 (58.3%) | 17 (54.8%) |  |
| Duration of flight (min) | 41.5 (20 - 155) |  | - |
| Peak ambient altitude (m) | 4267.2 (914.4 – 11887.2) |  | - |
| Max cabin altitude (m) | 899.2 (16.2 – 2286) |  | - |
| **Respiratory support** |  |  |  |
| Mechanical ventilation | 3 (12.5%) | 0 (0 %) | 0.15 |
| CPAP | 6 (25.0%) | 14 (45.2%) |  |
| Humified High Flow | 0 (0%) | 1 (3.2%) |  |
| Low flow | 4 (16.7%) | 3 (9.7%) |  |
| No support | 11 (45.8%) | 13 (41.9%) |  |
| Maximum FiO<sub>2</sub> | 0.23 (0.21 – 0.4) | 0.25 (0.21 – 0.3) | 0.83 |
| **Physiology parameters** |  |  |  |
| Mean BP (mmHg) | 50 (42 – 54) | 53 (45 – 57) | 0.15 |
| Haemoglobin (g/L) | 187 (154 – 201) | 169 (146.5 – 194) | 0.23 |
| Peripheral oxygen saturation (SpO<sub>2</sub>) | 95.2 (92.6 - 97.7) | 94.8 (91.1 - 96.9) | 0.33 |

For infants transported by air, the mean (standard deviation; SD) cerebral oxygen crS02 was lower at altitude than during the ground transportation phase (75.3 (6.2) vs 77.9 (6.22); p=<0.0001). For infants transported by road, there was no difference in CrS02 between the 1^st^ and 3^st^ quarter of their journey (79.0 (4.5) vs. 79.6 (4.6); p=0.09). Cerebral mean (SD) FTOE was higher at altitude than during ground transportation (0.18 (0.07) vs. 0.21 (0.07); p=<0.001). There were no differences in the mean (SD) cerebral cFTOE of transported by road between the 1^st^ and 3^st^ quarter of their journey (0.16 (0.05) vs. (0.16 (0.05); p=0.82).

In infants transported by air the mean (SD) mrSO_2_ was similarly lower at altitude than during the ground transportation phase (69.3 (16.6) vs. 74.2 (11.6); p=<0.01). There were no differences in mean (SD) mrS02 in the road group 1^st^ and 3^st^ quarter of the road journey (76.9 (9.4) vs. 78.0 (9.2); p=0.18). Mean (SD) mesenteric FTOE (mFTOE) was higher at altitude than during ground transportation (0.22 (0.12) vs. 0.28 (0.17); P=0.006). The mean (SD) mFTOE did not differ during road transit in the 1^st^ and 3^st^ quarter of the journey (0.19 (0.1) vs 0.18 (0.1); p=0.75) (Figure 1).

**Figure 1.**
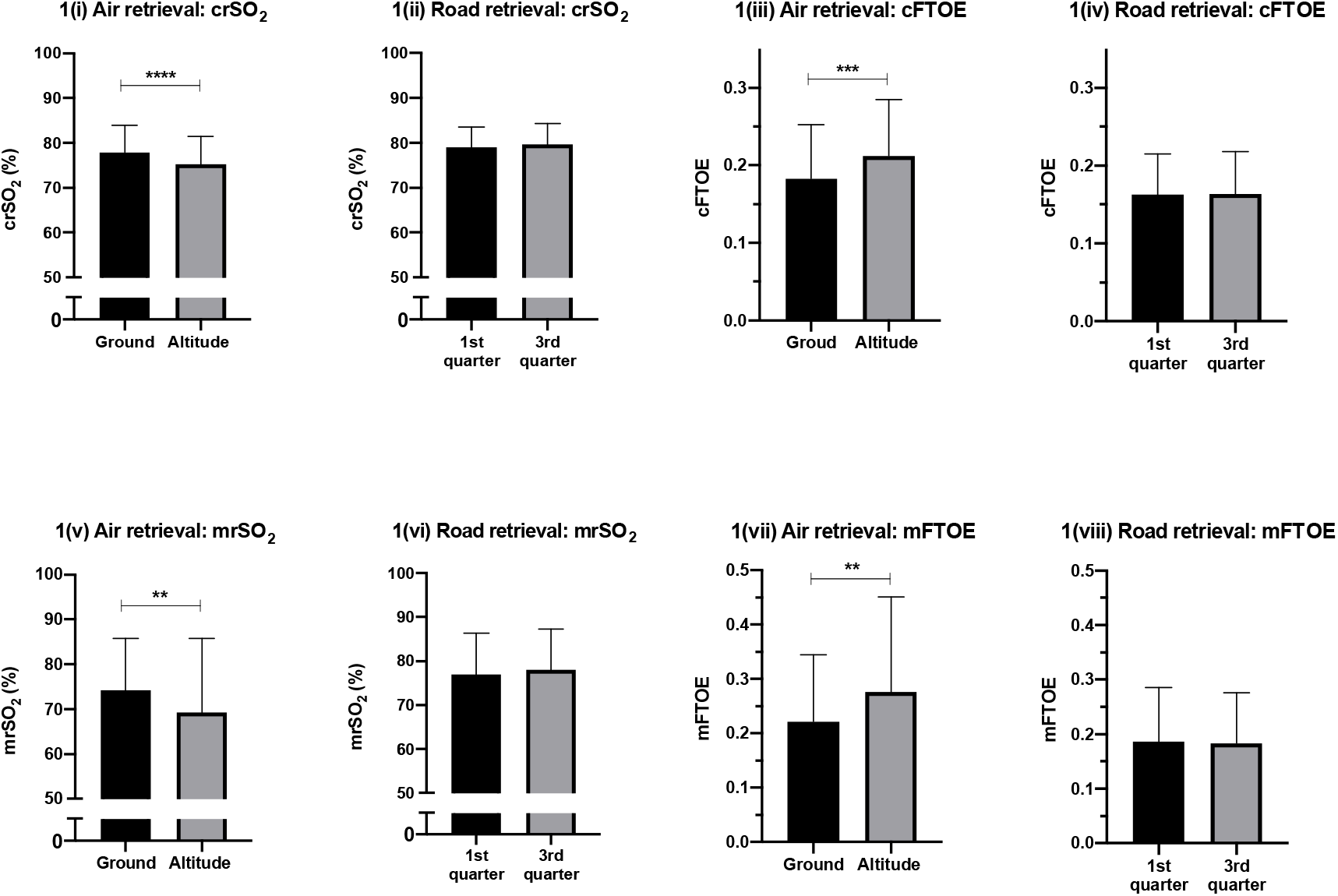
Graph of oxygenation (NIRS; crS02 and mS02) readings and Cerebral and Mesenteric Fractional Tissue Extraction (cFTOE and mFTOE) in infants transported at altitude and by road. Depicts NIRS Cerebral Oxygen Saturation (crS02), Mesenteric Oxygen Saturation Readings (mrS02), Cerebral Fractional Tissue Extraction (cFTOE) and Mesenteric Fractional Tissue Extraction (mFTOE). The error bars indicate mean with standard deviation. Figure 1(i) X axis displays the ground and altitude comparison columns, with Y-axis depicting cerebral NIRS percentage, p-value = <0.0001. Figure 1(ii) X-axis depicts comparison between first and third quarter of road transportation, with Y-axis the cerebral NIRS percentage, p-value =0.09. Figure 1(iii) X-axis is ground and altitude comparison, with Y-axis depicting cerebral fractional tissue extraction, p value = <0.001. Figure 1(iv) The X-axis depicts comparison between first and third quarter of road transportation and Y-axis the cerebral fractional tissue extraction, p value = 0.82. Figure 1(v) The X-axis depicts the ground and altitude comparison columns, with Y-axis depicting mesenteric NIRS percentage, p value =<0.01. Figure 1(vi) X-axis depicts comparison between first and third quarter of road transportation, with Y-axis the mesenteric NIRS percentage, p-value =0.18. Figure 1(vii) X axis is ground and altitude comparison, with Y-axis depicting mesenteric fractional tissue extraction, p value = <0.01. Figure 1(viii) The X-axis depicts comparison between first and third quarter of road transportation, with Y-axis the mesenteric fractional tissue extraction, p value=0.75.

## Discussion

Infants transported by air soon after birth in Western Australia are most likely to be retrieved around term gestation on the first day of life with respiratory disease. In these babies cerebral and mesenteric oxygenation were lower at peak altitude and oxygen extraction increased. There was no change in regional oxygenation or extraction in infants transported only by road.

There is little published data describing NIRS monitoring in the neonatal transport setting. Stroud et al^5^ examined cerebral oxygenation data at baseline and at cruising altitude on 17 paediatric patients during helicopter transportation (unpressurised environment) and similarly to our study observed that patients transported at > 1524 m (5000 feet) experienced a significant difference in NIRS readings These changes in cerebral oximetry were not accompanied by similar decrease in pulse oximetry. Hamrin et al^6^ demonstrated that in 44 children transported by fixed-wing twin engine turboprop, cerebral and splanchnic oxygen saturation decreased at altitudes of >1524 m (5000 feet).

In our study peripheral arterial saturations were similar at altitude and on the ground despite the differences in NIRS readings. Pulse oximetry is standard practice to ensure patients are receiving adequate supplemental oxygen during neonatal retrieval. NIRS-measured regional oxygenation reflects the balance of local tissue oxygen supply and demand while pulse oximetry reflects only on oxygen supply to tissue. Tobias et al^7^ have demonstrated that rS02 detects alterations in oxygenation prior to oxygen saturation monitoring. In a study of 46 ex-preterm infants being transferred on commercial flights at near-term corrected age, 35% required in-flight oxygen for significant desaturations^8^. In an in-flight simulation, 34 infants less than one year were exposed to 15% oxygen for 7 h to mimic the relative hypoxic environment of flight^9^. Sixty-two percent of the had irregular breathing patterns, periodic apnoeas, and significant desaturations.

There is more published data describing NIRS monitoring in the NICU. Evidence to support the regular use of NIRS and the impact on clinical outcome remains unclear. In a prospective observational cohort (n=734) infants less than 32 weeks’ gestation by Alderliesten et al^10^ showed that cerebral hypoxia in the first 72hrs of life was associated with unfavourable cognitive outcomes at 24 months. Alderliesten et al^11^ also published a case-control study of 132 preterm infants demonstrating that early cerebral hypoxia burden was associated with impaired neurodevelopmental outcomes while hypotension requiring inotropic support was not. The multi-centre feasibility Safe-BoosC phase II trial RCT (n=166) found that NIRS monitoring combined with a pragmatic treatment algorithm reduced cerebral hypoxia and hyperoxia burdens in preterm infants^12^. The European Society of Paediatric Research Special Interest Group on NIRS currently recommends the continued used of NIRS as an adjunctive tool rather than a tool to guide interventions^13^. The now completed Safe-BoosC phase III^14^ trial, (<u>NCT03770741</u>) analysed the benefit and harms of protocolized responses to NIRS, by acting on keeping the cerebral NIRS over the 10^th^ centile for the first 72 hours and results are awaited with interest.

Limitations to our prospective observational study are the small sample size of 55 infants with some heterogeneity in pathology requiring transport for tertiary care. Both the underlying aetiology of their disease process and their gestation may alter their haemodynamics and impact NIRS measurements. There is also a disproportionate number of males at 70% of the studied population. The potential impact of vibration and noise signal during air transport on the NIRS readings may further limit interpretation.

We have demonstrated in this small observation study that air transport reduces cerebral and mesenteric oxygenation and increases tissue oxygen extraction in transported soon after birth. Further study in a larger and more diverse population including preterms, in infants undergoing postnatal adaption without a disease process and more balanced male/female population are required to examine other factors which may have an impact on tissue oxygenation and extraction in newborns. The impact of lower cerebral oxygenation on the developing brain and its long-term affect requires longitudinal investigation.

## Data Availability

All data produced in the present work are contained in the manuscript

## Acknowledgments

We would like to acknowledge the families who participated in this research study

## Contribution Statement

All authors made a substantial contributions to the conception or design of the work and were involved in drafting the work or revising it critically for important intellectual content and gave final approval of the version to be published.

## References

1. Holt J, Fagerli I. Air transport of the sick newborn infant: audit from a sparsely populated county in Norway. Acta Paediatrica 2007;88(1):66–71. doi: 10.1111/j.1651-2227.1999.tb01271.x

2. Samuels MP. The effects of flight and altitude. Arch Dis Child 2004;89(5):448–55. doi: 10.1136/adc.2003.031708 [published Online First: 2004/04/23]

3. JD Tobias. Cerebral oximetry monitoring with near infrared spectroscopy detects alterations in oxygenation before pulse oximetry. J Intensive Care Med 2008;Nov-Dec 2008, 23(6):384–8. doi: 10.1177/0885066608324380.

4. Bel F V. Monitoring Neonatal Regional Cerebral Oxygen Saturation in Clinical Practice: Value and Pitfalls,. Neonatology 2008;94:237–244

5. Michael H Stroud, Punkaj Gupta, Parthak Prodhan. Effect of altitude on cerebral oxygenation during pediatric interfacility transport. Pediatr Emerg Care 2012;28(4):329–32. doi: 10.1097/PEC.0b013e31824d8b3c

6. Hannegård Hamrin T Es, Berner J, Fläring U et al. Influence of altitude on cerebral and splanchnic oxygen saturation in critically ill children during air ambulance transport. PLoS One 2020;15(9):e0239272.

7. Tobias JD. Cerebral oximetry monitoring with near infrared spectroscopy detects alterations in oxygenation before pulse oximetry. J Intensive Care Med Nov-Dec 2008;23(6):384-8.;Nov-Dec 2008.(23(6):384-8.) doi: 10.1177/0885066608324380.

8. Resnick Steven M. F Hall Graham L. P Simmer Karen N., fPhd, et al. The Hypoxia Challenge Test Does Not Accurately Predict Hypoxia in Flight in Ex-Preterm Neonates. Chest Journal 2008;133(5):1161–66.

9. Parkins KJ, Poets CF, O’Brien LM, et al. Effect of exposure to 15% oxygen on breathing patterns and oxygen saturation in infants: interventional study. BMJ (Clinical research ed) 1998;316(7135):887–91. doi: 10.1136/bmj.316.7135.887 [published Online First: 1998/04/29]

10. Alderliesten T. Low Cerebral Oxygenation in Preterm Infants Is Associated with Adverse Neurodevelopmental Outcome. The Journal of Pediatrics,; Volume 207,: 109-16.e2.

11. Thomas Alderliesten M, Petra M.A. Lemmers, MD, PhD, Ingrid C. van Haastert, MA, PhD, Hypotension in Preterm Neonates: Low Blood Pressure Alone Does Not Affect Neurodevelopmental Outcome. The Journal of Pediatrics, 2014;154(5):986–91.

12. Plomgaard AM, van Oeveren W, Petersen TH, et al. The SafeBoosC II randomized trial: treatment guided by near-infrared spectroscopy reduces cerebral hypoxia without changing early biomarkers of brain injury. Pediatric Research 2016;79(4):528–35. doi: 10.1038/pr.2015.266

13. Mathias Lühr Hansen, SH-S, Janus, Christian Jakobsen et al, European Society for Paediatric Research Special Interest Group ‘NearInfraRed Spectroscopy’ (NIRS), Collaborators A. Cerebral near-infrared spectroscopy monitoring (NIRS) in children and adults: a systematic review with meta-analysis. Pediatr Res 2022 2022 doi: doi: 10.1038/s41390-022-01995-z

14. Hansen ML, Pellicer A, Gluud C, et al. Cerebral near-infrared spectroscopy monitoring versus treatment as usual for extremely preterm infants: a protocol for the SafeBoosC randomised clinical phase III trial. Trials 2019;20(1):811. doi: 10.1186/s13063-019-3955-6 [published Online First: 2020/01/01]

